# Determinants of SARS-CoV-2 anti-spike antibody levels following BNT162b2 vaccination: cross-sectional analysis of 6,000 SIREN study participants

**DOI:** 10.1101/2022.04.21.22274025

**Authors:** Ashley David Otter, Silvia D’Arcangelo, Heather Whitaker, Jacqueline Hewson, Sarah Foulkes, Ana Atti, Michelle Cole, Ezra Linley, Simon Tonge, Nipunadi Hettiarachchi, Noshin Sajedi, Davina Calbraith, Chris Norman, Elen de Lacy, Lesley Price, Sally Stewart, Lisa Cromey, Diane Corrigan, SIREN study group, Cathy Rowe, Colin S Brown, Jasmin Islam, Amanda Semper, Susan Hopkins, Victoria Hall, Tim Brooks

**Author notes:** **Corresponding author:** Ashley Otter. SIREN study group authors listed within the supplementary material.

## Abstract

**Background:** Understanding immunological responses to SARS-CoV-2 vaccinations is integral to the management of SARS-CoV-2. We aimed to investigate determinants of antibody response to the BNT162b2 vaccine.

**Methods:** A cross-sectional analysis of anti-spike binding antibodies in serum samples from healthcare workers after one or two doses. Post-vaccination interval was restricted to ≥21 days after dose 1, ≥14 days after dose 2. The primary outcome was anti-S titres with explanatory variables dose, previous infection, dosing interval, age, ethnicity, and comorbidities. Multivariable linear regression was also conducted.

**Results:** Participants (n=5,871) included 3,989 post-dose 1, 1,882 post-dose 2. In SARS-CoV-2 infection naïve, 99.65% seroconverted after dose 1 and >99.9% seroconverted after dose 2. Geometric mean anti-S titre in the naïve cohort was 75.48 Binding Antibody Units/ml after dose 1, 7,049 BAU/ml after dose 2. Anti-S titres were higher in those with previous infection (2,111 BAU/ml post-dose 1, 16,052 BAU/ml post-dose 2), and increased with time between infection and vaccination: 3 months 1,970 (1,506-2,579) vs 9 months; 13,759 (8,097-23,379). Longer dosing intervals increased antibody response post-dose 2: 11-fold higher with a longer interval (>10 weeks) than those with shorter intervals, across all age-groups. Younger participants had higher mean titres (>2.2-fold higher). Multivariable regression modelling corroborated the above associations, and also found higher titres associated with being female or from an ethnic minority but lower titres among immunocompromised participants.

**Conclusion:** The number of antigen exposures and timing between vaccinations plays a significant role in the magnitude of the post-vaccination antibody response, with implications for long-term protection and post-booster antibody responses.

## Introduction

Since its emergence in December 2019, SARS-CoV-2 is the largest respiratory virus pandemic this century, resulting in around 505 million COVID-19 cases and over 6.2 million deaths^1^, including 171,396 within the UK^2^. Towards the end of 2020, vaccines against SARS-CoV-2 became available, including the mRNA vaccines (BNT162b2 (Pfizer/BioNTech) and mRNA-1273 (Moderna)), and the chimpanzee adenovirus vectored-vaccine ChAdOx-1 nCoV-19 AZD1222 (Oxford/Astra Zeneca). Whilst single-dose vaccinations have provided some protection from disease in animal models^3^ and have demonstrated robust antibody responses^4–6^, data from phase 3 clinical trials of all these vaccines showed highly efficacious protection against severe disease following two vaccine doses, with trial participants demonstrating robust and sustained immunity^6,7^.

On the 2^nd^ December 2020, the MHRA granted temporary authorisation for the use of the Pfizer/BioNTech mRNA vaccine (BNT162b2, Comirnaty) for use within the UK^8^ and the first doses were given on the 8^th^ December 2020^9^. Care home residents, those over 80 years of age and frontline health and social care workers were the primary focus of the initial SARS-CoV-2 vaccine rollout in the UK, with frontline healthcare workers first being offered the BNT162b2 vaccine in December 2020. In line with the evidence from the manufacturer’s clinical trials, the vaccine programme initially recommended 3 weeks between the first and second doses of BNT162b2^7^. Faced with a finite supply of vaccines, in January 2021 the Joint Committee on Vaccination and Immunization (JCVI) recommended extending the interval between first and second dose to up to 12 weeks for both the Pfizer and Oxford Astra/Zeneca vaccines, in order to maximise the number of people receiving a first dose and therefore some level of protection against SARS-CoV-2^10,11^.

As of March 2022, 86.2% of the UK population aged over 12 years had received at least two doses, whilst 92.1% had been given at least one dose of a SARS-CoV-2 vaccine^12^. With increases in disease prevalence and the spread of variants of concern (e.g. Delta and Omicron), the vaccines have shown their capability to prevent and protect against severe SARS-CoV-2 disease, but further work is ongoing to understand durability of the immune response, correlates of protection and why some individuals develop COVID-19 even when double vaccinated.

The SARS-CoV-2 Infection and Reinfection EvaluatioN (SIREN) is a longitudinal study of over 44,000 healthcare workers across the UK^13–15^. Its initial focus was to assess the impact of infection/reinfection on immunity, but it has since been applied to study vaccination responses in detail. The aim of this paper is to analyse the levels of anti-spike binding antibodies and after COVID-19 vaccination and/or SARS-CoV-2 infection, stratified by age, previous infection, gender, health conditions, vaccine dose, timing between sample collection and vaccination, and dosing interval.

## Methods

### Study design and setting

For this cross-sectional analysis, a convenience sample was drawn from the cohort of healthcare workers enrolled within the SARS-CoV-2 Infection and Reinfection EvaluatioN (SIREN) study^13–15^. Inclusion and exclusion criteria for the main analysis can be found in the supplementary material (**Supplementary figure 1**). In brief, eligible participants had submitted an enrolment sample (baseline sample) that had been tested using the Roche Elecsys quantitative SARS-CoV-2 anti-spike (S) and semi-quantitative anti-nucleocapsid (N) antibody assays and submitted this after their first or second COVID-19 vaccination, or were anti-N positive and submitted the sample before vaccination. Only participants receiving 1 or 2 doses of BNT162b2 were included, with vaccines other than BNT162b2 or unknown vaccines excluded due to low vaccination numbers (e.g. ChAdOx-1, mRNA-1273). To prevent bias in the data from participants still mounting an antibody response at the time of sampling, only those participants that gave a baseline sample ≥21 days post dose 1 or post-infection, and those ≥14 days post dose 2 were included in the final analysis.

### Data sources

Vaccination data (manufacturer, dates) were obtained via linkage on personal identifiers from national COVID-19 vaccination registries and directly from participants in their study questionnaires. Site test results reported into national laboratory surveillance systems (in England the Second-Generation Surveillance System (SGSS), with equivalent data securely transferred from Northern Ireland, Wales and Scotland) were linked to individuals’ personal information provided in the enrolment questionnaire, with testing data updated daily and captured in a SQL SIREN Database. Serology results from standardised antibody testing at the UKHSA SARS-CoV-2 Serosurveillance laboratory were captured in the SIREN database through linkage on unique identifiers. Data from participants with baseline serum samples with confirmatory antibody test results were extracted from the SIREN SQL database.

### Outcomes

The primary outcome was anti-S titre using the Roche Elecsys anti-spike (ACOV2 S) assay which specifically targets anti-RBD antibodies, as described elsewhere^4,16^. Samples were considered positive for anti-nucleocapsid and anti-spike antibodies if the results were ≥1.0 COI and ≥0.8 U/ml, respectively. The Roche Elecsys anti-nucleocapsid assay is semi-quantitative, as described elsewhere^17^, with the presence of anti-nucleocapsid antibodies used as a proxy for probable previous infection in the absence of a previous PCR positive. The Roche anti-spike assay is quantitative, producing results between 0.4 U/ml and 225,000 U/ml, with automatic dilutions performed to achieve sample results within the quantitative range (0.4 to 250 U/ml). The Roche anti-spike assay has been fully calibrated to the NIBSC first WHO International SARS-CoV-2 immunoglobulin standard (NIBSC code 20/136), enabling reporting in binding antibody units (BAU) per ml (BAU/ml), with all analyses here presented as BAU/ml^18^.

### Explanatory variables

Participants were characterised according to their previous infection status: ‘naïve’ (anti-nucleocapsid negative at baseline and no linked PCR positive results), ‘previous infection with PCR+’ (anti-N positive at baseline and linked previous SARS-CoV-2 PCR positive result), ‘previous infection without PCR+’ (no known previous SARS-CoV-2 PCR positive date but presence of anti-nucleocapsid antibodies). Samples from cohorts of participants pre-vaccination and post-vaccination were used to compare post-vaccination and post-infection antibody titres.

Outcomes were stratified by dosing interval, age, gender, ethnicity and comorbidities. For analysis on existing health conditions, participants were classified into three categories based on data provided within their enrolment questionnaire. Specific health conditions were categorised: immunosuppression (participants with immune system cancer, rheumatology disease, transplant recipients, those with spleen conditions or other autoimmune conditions not listed here), chronic respiratory disease (asthma, chronic obstructive pulmonary disease, or other chronic respiratory diseases), and chronic non-respiratory diseases (diabetes, obesity, chronic-neurological disease, dementia, other cancers, chronic heart disease, chronic kidney disease, liver disease and HIV).

### Statistical analysis

All statistics were performed on log transformed anti-spike antibody values (in BAU/ml), unless otherwise stated, using R (version 4.0.2), with regression analysis performed in STATA (version 14.2). Mean antibody titres are reported as geometric means. Two-sample t-tests were used for comparisons between groups. Normal error multivariable linear regression models were fitted to log antibody levels (separate models for 28+ days post dose 1 and 21+ days post dose 2), including covariates such as days since dose, age group, sex, ethnicity, existing health conditions and for dose 2 only, dosing schedule group. Regression coefficients were exponentiated for interpretation as adjusted geometric mean ratios. The interval in days between vaccine dose and the baseline SIREN sample was modelled as log linear (i.e. exponential decay on the original scale). All other covariates were categorical.

### Ethics

Ethics approval was granted on 22 May 2020 under IRAS ID 284460, Berkshire Research Ethics Committee Approval by HRA and Health and Care Research Wales.

## Results

A total of 13,002 participants were included in this analysis, 5,871 post vaccination; ≥21 days post-dose 1 (n=3,989) and ≥14 days post-dose 2 (n=1,882), and 7,131 participants who provided baseline samples prior to vaccination and had evidence of previous infection. Most participants were female (82.25%) and of white ethnicity (86.94%) and were distributed across the different regions of England (n=9,007, 69.27%) as well as Wales (n=669, 5.15%), Scotland (n=2,860, 22.00%), and Northern Ireland (n=466, 3.58%).

Cohort assignment was as follows: naïve cohort (post-dose 1: n=2,863; post-dose 2: n=1,523); previous infection with PCR+ (post-dose 1: n=571; post-dose 2; n=165); previous infection without PCR+ (post-dose 1: n=555; post-dose 2, n=194). Further classification and numbers per infection cohort, gender and ethnicity can be found in **Table 1** (with breakdown further shown in supplementary table 1). Within the pre-vaccination cohort, participants were similarly assigned previous infection with PCR+ (n=1,990) or previous infection without PCR+ (n=5,141).

**Table 1:**
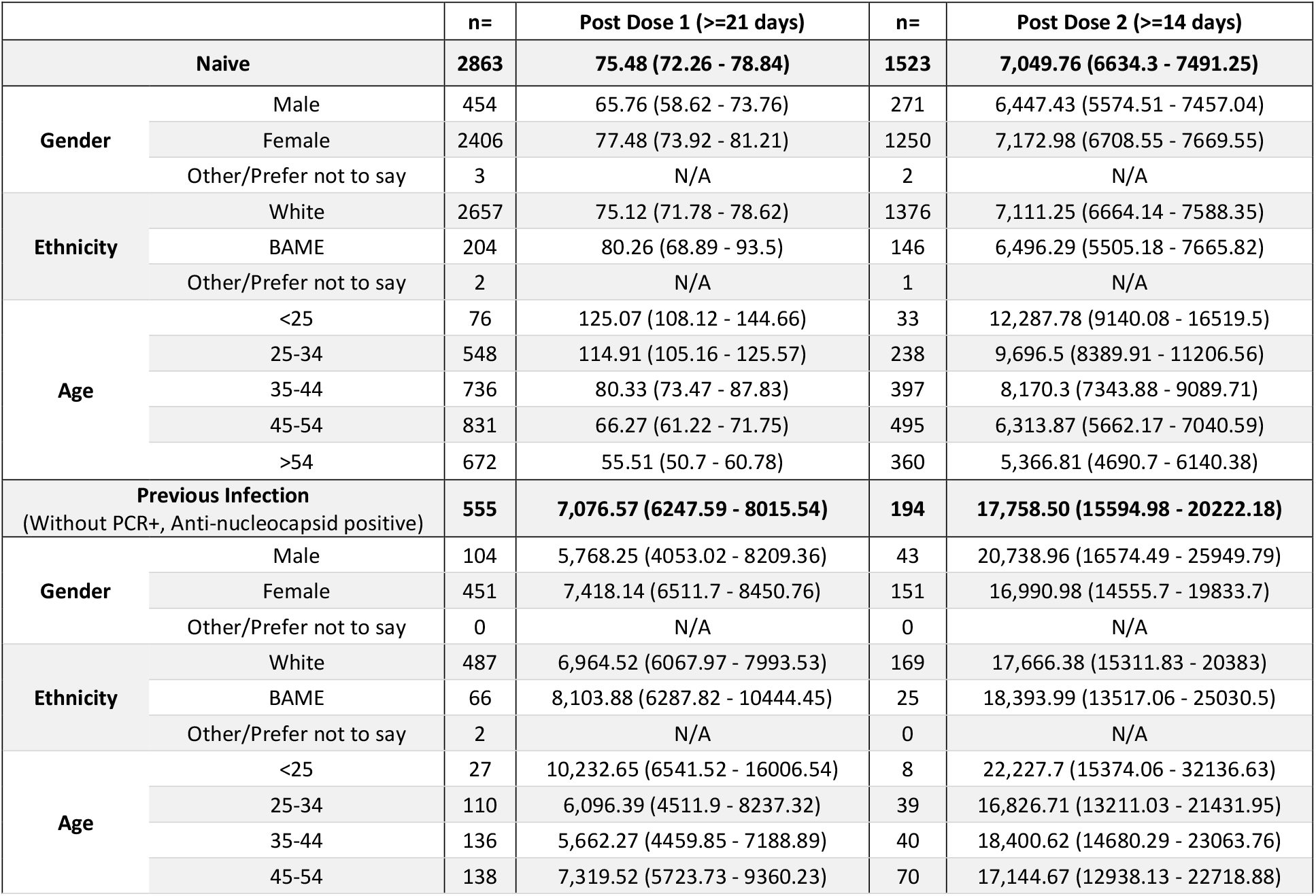
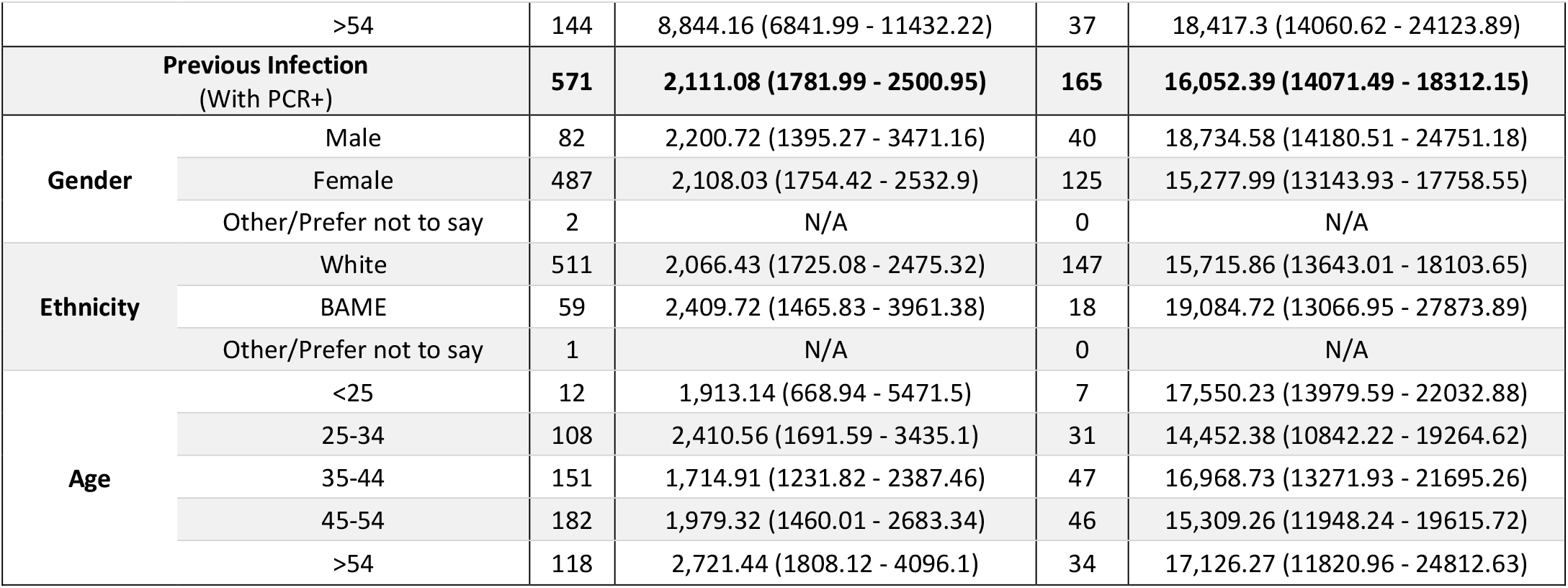
Anti-spike antibody titres for the post-vaccination SIREN cohort. All data is split according to prior infection status (naïve, previous infection without PCR+, or previous infection with PCR+) as well as gender, ethnicity and age. Data are presented as geometric means of the Roche S values (BAU/ml) with 95% CIs. Those categories with less than 5 participants were removed due to high statistical variance.

In the naïve cohort 2,853 participants (99.65%) seroconverted after dose 1, with detectable anti-spike antibodies (≥0.8 BAU/ml). Of the 10 participants that did not seroconvert post-dose 1 (0.35%), the median age was 51 years (Range 40 – 65, IQR 44.5 – 56.75) and a median of 41 days between first vaccination and sample date (Range 24 – 71, IQR 36.25 – 46.75). When analysing the sera of participants ≥14 days post-dose 2, 1,522/1,523 participants seroconverted (99.93%), with only one participant aged in their 50s not seroconverting within 29 days between dose 2 and the sample date.

### Previously infected individuals mount a significantly higher anti-spike antibody response post-dose 1 and post-dose 2

Geometric mean levels of anti-spike binding antibody titres post-dose 1 from all naïve participants (n=2,863) were 75.48 BAU/ml (95% CI: 72.26 –78.84), whereas significantly higher titres were measured in those with previous infection with PCR+ (n=571, 2,111 BAU/ml (1,782 – 2,501); p=<0.0001) and previous infections without PCR+ (n=555, 7,077 BAU/ml (6,248 – 8,016); (p=<0.0001) (Supplementary Figure 2).

Geometric mean titres post-dose 2 from all naïve participants (n=1,523) were 7,050 BAU/ml (6,634 – 7,491), whilst those with previous infection with PCR+ (n=165) and previous infection without PCR+ (n=194) were significantly higher at 16,052 BAU/ml (14,071 – 18,312, p=<0.0001) and 17,759 BAU/ml, respectively (15,595 – 20,222, p=<0.0001) (Supplementary Figure 2).

Previous infection results in comparable antibody titres to 1 vaccine dose. In those with previous infection with PCR+, geometric mean antibody titres post-infection were 96.99 BAU/ml (89.53 – 105) whilst those with previous infection without PCR+ had a geometric mean titre of 81.20 (77.50 – 85.07), which is highly similar to the naïve group receiving their first dose, with a geometric mean titre of 75.48 (72.26 –78.84).

### Age and dosing interval have significant effects on post-vaccination antibody titres

Naïve older participants had significantly lower antibody responses post-dose 1 than younger HCWs (**Figure 2**); there was a ∼2-fold decrease between the 25-34 years (115 BAU/ml, 95% CI 105 - 126) and >54 years (55.51 BAU/ml, 95% CI 50.7 - 60.78) age groups (2.03, 95% CI 1.78-2.32). Similarly, a ∼2.3-fold decrease was observed between the 25-34 (12,288 BAU/ml; 95% CI 9,140 – 16,520) and >54 years (5,367 BAU/ml, 95% CI 4,691 – 6,140) age groups post-dose 2 (1.63, 95% CI 1.34-1.98). In contrast, those with previous infection, showed no significant differences between any of the age groups post-dose 1 or post-dose 2 (Figure 1).

**Figure 1:**
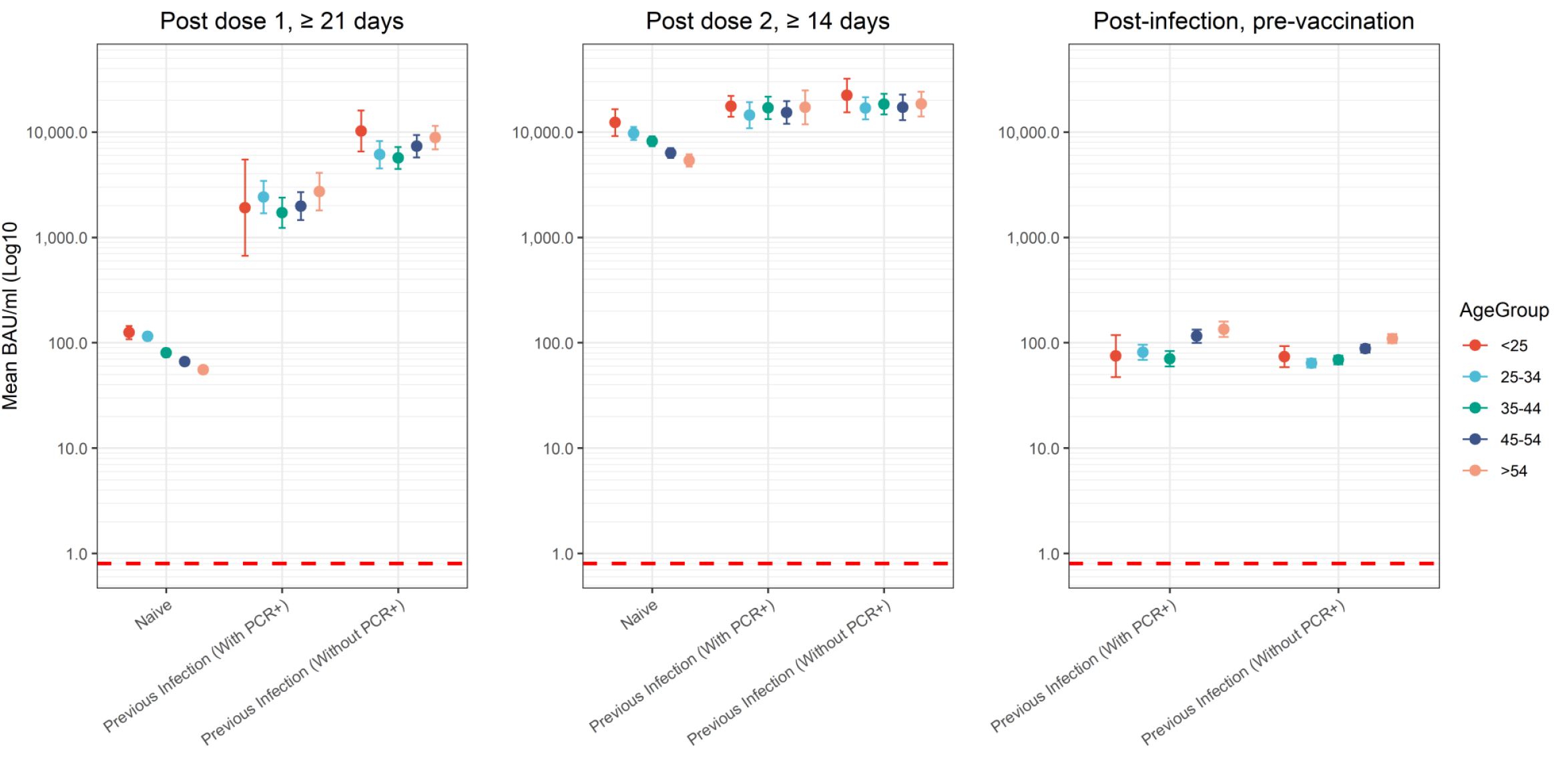
Antibody levels post-dose 1 (left), post-dose 2 (middle) and post-infection (right), split by cohort and age group. Naive - those with no known previous COVID-19 disease (no PCR confirmation or anti-nucleocapsid antibodies at baseline). Previous infection with PCR+ – those with a confirmed PCR positive. Previous infection without PCR+, those with no known PCR positive date, but presence of anti-nucleocapsid antibodies.

**Figure 2:**
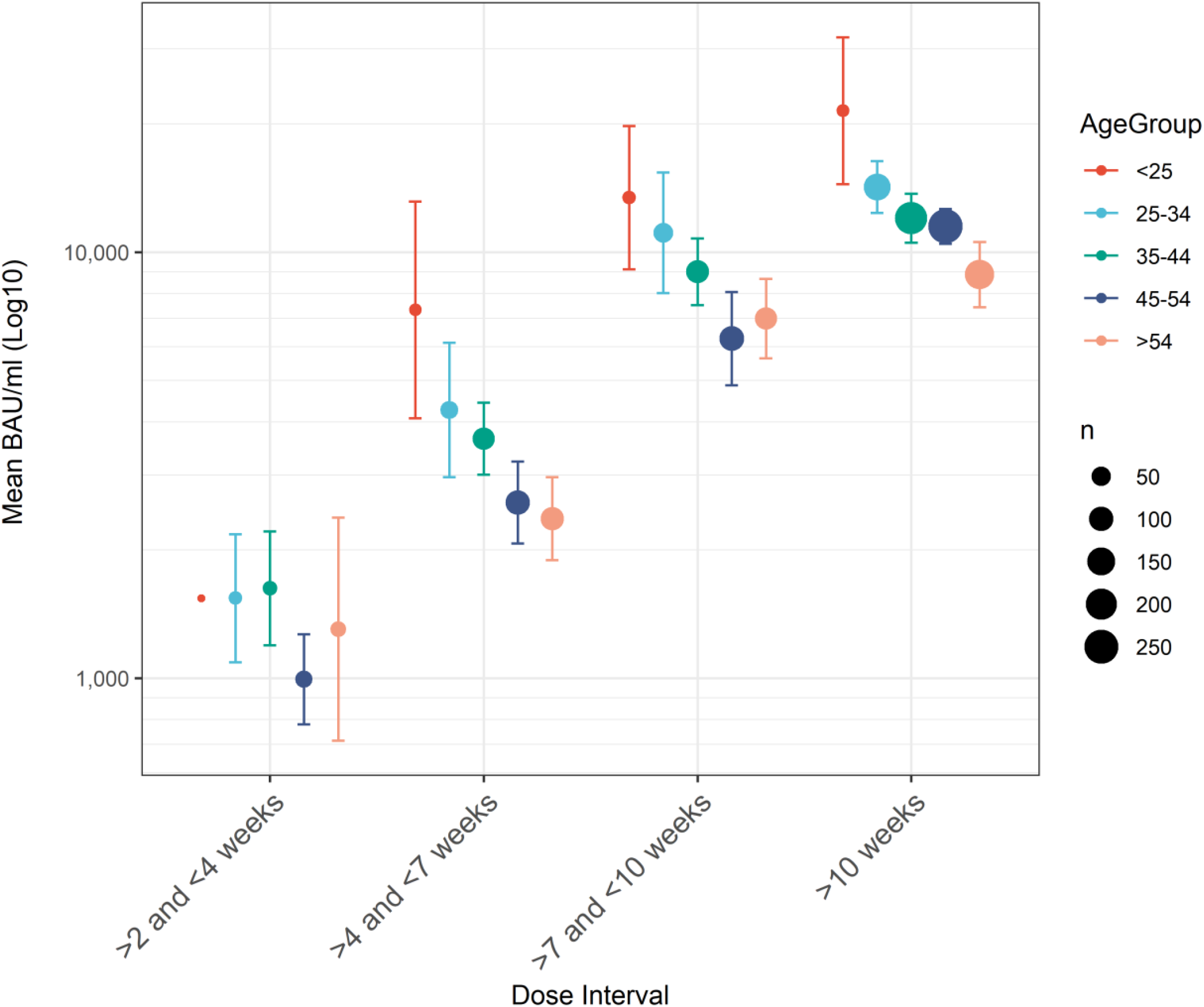
Geometric mean anti-spike antibody levels (BAU/ml) of naïve participants post dose 2 split according to dosing interval and age. Mean BAU/ml increases with dosing interval but decreases with age. Only 1 participant was available within the <25 age category and 2-3-week interval group. The circle diameters correspond to the number of participants for each age group at each dosing interval. Error bars shown are 95% CI.

The geometric mean antibody titres increased with longer intervals between doses, with a 9-fold increase after an interval of >10 weeks compared with >2 and <4 weeks, with this trend consistent across all age groups (Table 2). In general, younger participants had significantly higher antibody responses than older participants with the same vaccine interval, with the exception of the >2 and <4-week dose interval (due to low numbers in the <25 and 25-34 age groups) (Table 2). Thus, participants aged >54 years with a dosing interval of >2 and <4 weeks had a geometric mean antibody titre of 1,303 BAU/ml (n= 25, 95% CI 713.28 - 2382), whereas those aged <25 years with a >10 week dosing interval had a geometric mean antibody titre of 21,472 BAU/ml (n= 10, 95% CI 14,435 – 31,940), a ∼16.5 fold difference (Table 2). Antibody titres for each of the dosing interval groups were also analysed in relation to the time elapsed between vaccination and sample date, to prevent the data being skewed by sampling bias (Supplementary Table 3 and Supplementary figure 4). Irrespective of whether participants provided a baseline sample 4-5 weeks, 6-7 weeks or 8-9 weeks after their second dose, the significant difference between >2 and <4 weeks and >10 weeks dosing intervals remained, highlighting that the observed difference in antibody titres was due to the dosing interval and not due to sample collection bias.

**Table 2:** Anti-spike antibody titres (BAU/ml) of naive participants post-second vaccine dose, split according to age. Data are presented as geometric means, with 95% CIs. All groups exhibit higher anti-spike antibody titres with increasing interval, whilst decreasing titres are found with age group in all dosing intervals. GMC: Geometric mean (BAU/ml).

|  |  | Dose interval (time between 1<sup>st</sup> dose and 2<sup>nd</sup> dose) |  |  |  |  |  |  |  |
| --- | --- | --- | --- | --- | --- | --- | --- | --- | --- |
|  |  | >2 and <4 weeks |  | >4 and <7 weeks |  | >7 and <10 weeks |  | >10 weeks |  |
| Age group |  | n= | GMC (95% CI) | n= | GMC (95% CI) | n= | GMC (95% CI) | n= | GMC (95% CI) |
|  | <25 | 1 | 1,540<br>(1540 - 1540) | 9 | 7,320.66<br>(4076.85 - 13145.47) | 13 | 13,430.99<br>(9117.29 - 19785.62) | 10 | 21,472.12<br>(14435.06 - 31939.72) |
|  | 25-34 | 14 | 1,541.01<br>(1089.67 - 2179.29) | 39 | 4,261.08<br>(2962.14 - 6129.62) | 52 | 11,100<br>(8010.84 - 15380.4) | 133 | 14,205.57<br>(12350.18 - 16339.69) |
|  | 35-44 | 19 | 1,626.8<br>(1196.19 - 2212.42) | 76 | 3,648.89<br>(3003.64 - 4432.75) | 85 | 8,994.43<br>(7515.83 - 10763.92) | 217 | 12,018.95<br>(10531.43 - 13716.58) |
|  | 45-54 | 34 | 993.47<br>(778.96 - 1267.07) | 101 | 2,584.21<br>(2071.11 - 3224.42) | 103 | 6,267.91<br>(4869.24 - 8068.36) | 257 | 11,489.2<br>(10455.48 - 12625.12) |
|  | >54 | 25 | 1,303.47<br>(713.28 - 2382) | 87 | 2,368.84<br>(1893.59 - 2963.37) | 75 | 6,984.68<br>(5632.33 - 8661.74) | 173 | 8,862.21<br>(7428 - 10573.33) |

### Increasing time between previous infection and dose 1 results in increased antibody titres

Those with previous infection without PCR+ had a significantly higher geometric mean (p=<0.0001) anti-spike antibody level post-dose 1 (7,077 BAU/ml) than those with previous infection with PCR+ (2,111 BAU/ml). When further stratifying the previous infection with PCR+ group by time between infection (median; 101 days) and vaccine dose 1, in general, increasing antibody titres were seen with increasing months after infection, up to 6 months and plateauing (3 months; 1,971 (1,506-2,579) vs 9 months; 13,759 (8,098-23,379), p=<0.0001, **Figure3**A). No increase in antibody titre was observed with increasing months between infection and dose 2 (**Figure3**C).

**Figure 3:**
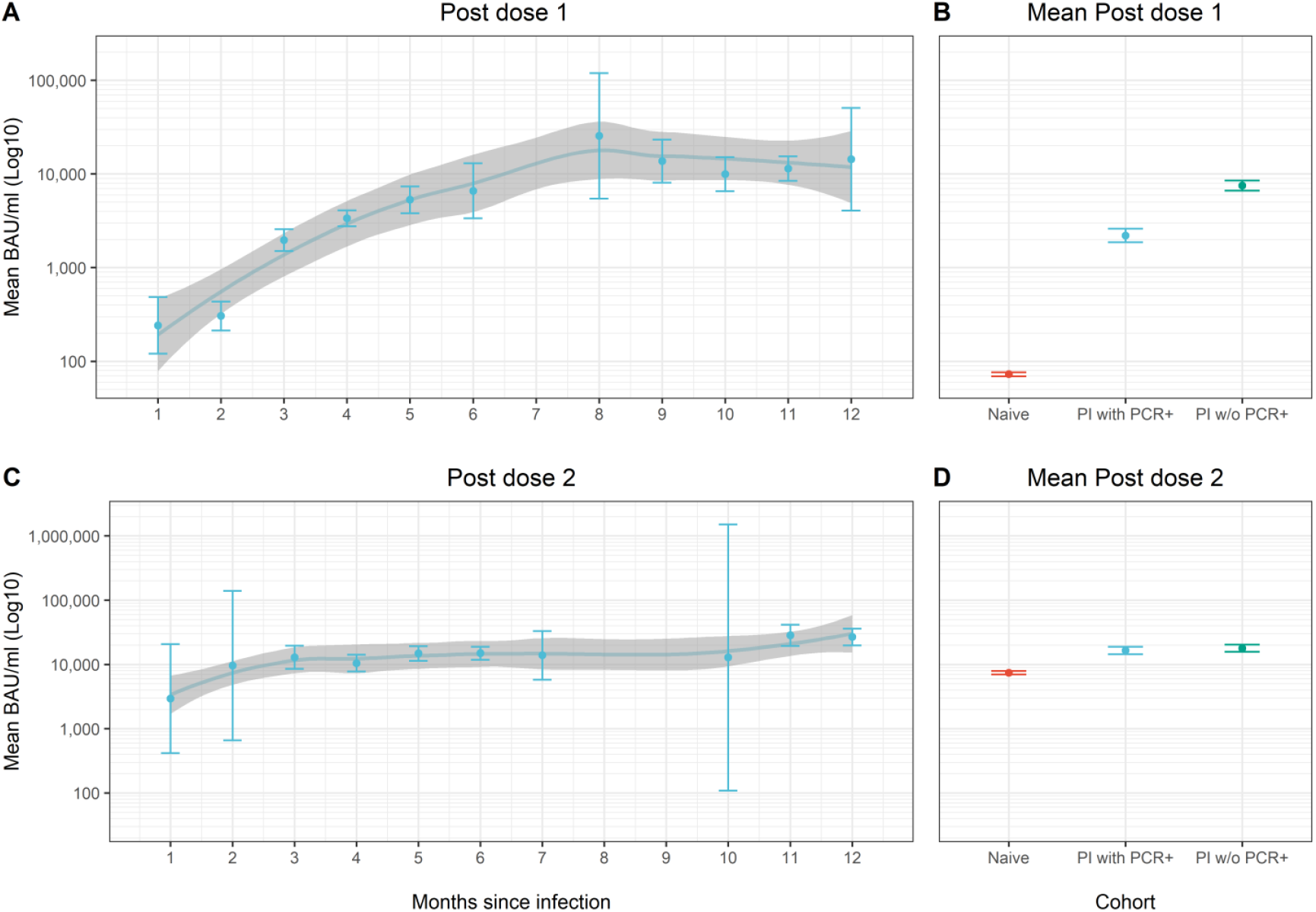
Post-dose 1 and post-dose 2 antibody titres in participants with previous infection (with history of PCR+), stratified by the interval (in months) between infection (PCR positive date) and vaccine dose 1 date (A) and vaccine dose 2 date (C). Mean post-dose 1 titres according to a participant’s exposure status post-dose 1 (B) and post-dose 2 (D). Data shows a general trend of increasing post-dose 1 antibody titres up to 6 months between infection and dose 1. Only participants that provided a baseline sample ≥21 days and <71 days (10 weeks) after dose 1, and those that provided a baseline sample ≥14 days and <71 days (10 weeks) after dose 2 were included. PI; previous infection, w/o; without.

### Multivariable regression models: the impacts of days since dose, age, gender, ethnicity, and prior health conditions on post-vaccination antibody levels

The decline in antibody levels following dose 1 was shallow, with a long half-life of 157.6 days (95% CI 92.1 - 546.2). Following dose 2 the decline was steeper, with a half-life of 45 days (95% CI 34.3 - 67.2). Adjusted geometric mean ratio of responses is shown (**Figure 4**, Supplementary table 2). Age was associated with antibody titres, with significantly higher titres in younger age groups. Female participants were found to have slightly higher antibody titres post-dose 1 (p=<0.001), but post-dose 2, this significance was not seen (p=0.151). Conversely, ethnic minority participants displayed no significant difference in antibody responses post-dose 1 (p=0.147), however, they did show a significantly higher antibody response post-dose 2 (p=0.024). No significant difference was observed between any of these groups post-dose 2 in those previously infected, with PCR+ or without PCR+.

**Figure 4:**
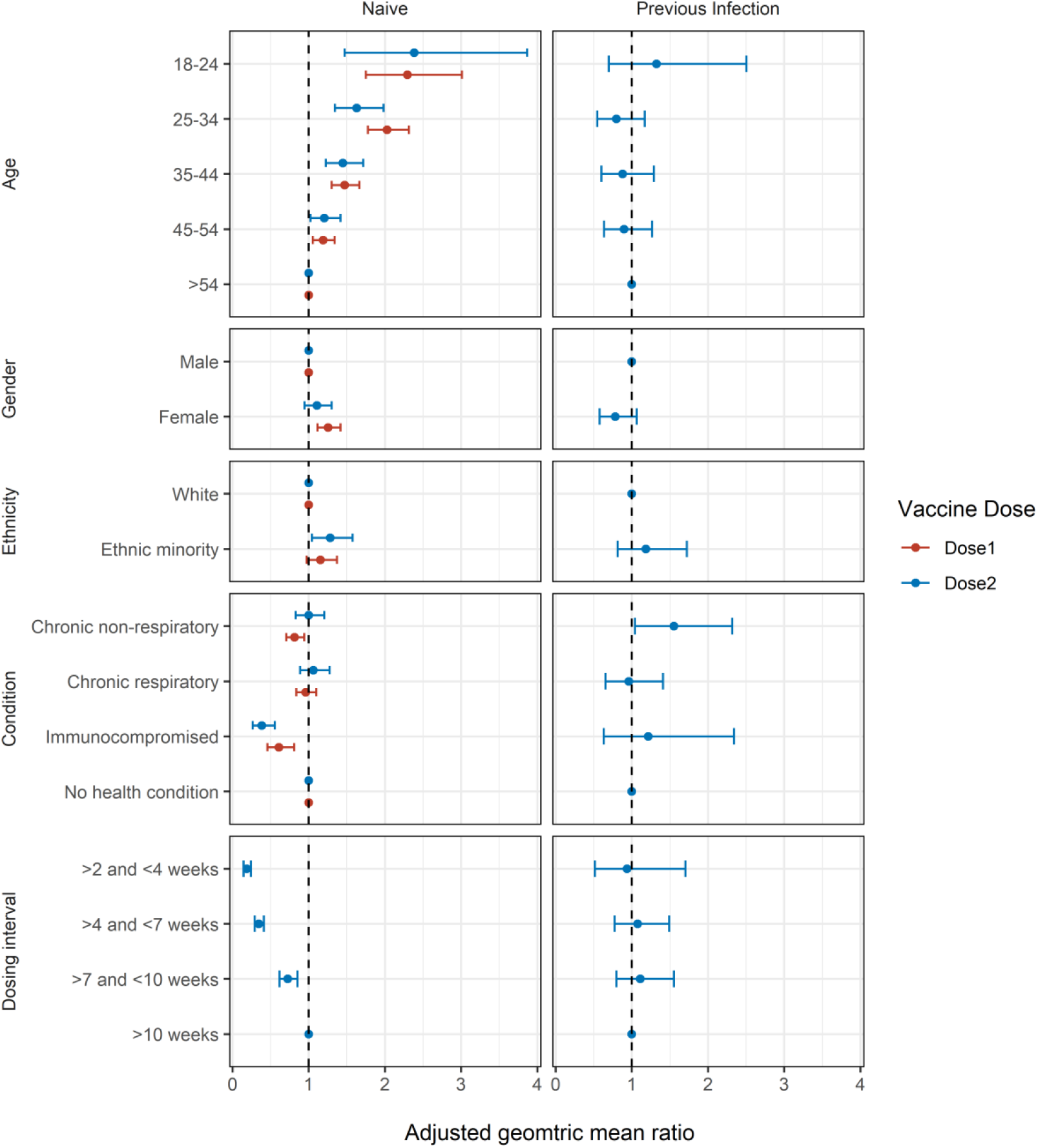
Multivariable regression analysis on anti-spike antibody levels, according to age group, dosing interval group, ethnicity, and gender, and also adjusted for days since dose. Data includes those post-dose-1 (≥28 days) and post-dose 2 (≥21 days). Age and dosing interval significantly affect antibody titres. Previous infection was grouped based on history of PCR+ or presence of anti-nucleocapsid antibodies. Due to low numbers, previous infected individuals with 1 vaccine dose were not included in this analysis

In naïve, immunosuppressed participants, antibody titres post-dose 1 and post-dose 2 were significantly lower than in participants with no immunosuppression (post-dose 1; p=0.001, post-dose 2 = p<0.001) However no significant difference was seen in those with previous infection post-dose 2 (p=0.563). Naïve participants with chronic non-respiratory diseases had a significantly lower antibody titre post-dose 1 (p=0.005), but no significant difference was observed in participants post-dose 2 (p=0.994). Chronic respiratory disease had no impact on post-dose 1 (p=0.566) or post-dose 2 (p=0.512) antibody titres, and similarly no difference was observed post-dose 2 in previously infected participants (p=0.830).

## Discussion

With data from 5,871 well-characterised participants after vaccination and 7,131 after infection, and all tested on the same anti-spike and anti-nucleocapsid assays, this dataset has provided a highly robust, cross-sectional analysis of determinants of the antibody response following vaccination. As this study used a fully quantitative anti-RBD assay reporting in BAU/ml relative to the 1^st^ WHO international SAR-CoV-2 immunoglobulin standard^19^, this enables meaningful comparison of post-vaccination responses in different cohorts and across diverse studies. Our data now offers a benchmark against which to compare results by age (and additional variables) across diverse post-vaccination studies, with comparisons to other major manufacturers available through the HARMONY study. We have shown that antibody response increases with repeated antigenic exposure, with participants with three antigen exposures (infection and then two vaccine doses) having the highest anti-S titres. We observed similar anti-S responses in those with a single antigen exposure, irrespective of whether this was from infection or vaccination. Anti-S titres were much higher after two vaccine doses than infection alone. Timing of second antigen exposure impacted antibody responses in our participants, with longer intervals between successive vaccine doses, or first vaccine dose after infection, associated with an increased antibody response as demonstrated by others^6,20–23^. We also found associations with age and immunosuppression, with higher anti-S titres in younger participants and lower titres in those with immunosuppression.

Other studies have also found higher anti-S titres following two antigen exposures, whether vaccination alone or infection and then a single vaccine dose^4,24–26^, but few have shown that those with prior infection continue to have higher titres than double-vaccinated infection-naïve individuals, suggesting that those with prior infection still benefit from vaccination. This finding of incremental increases in anti-S titre with repeated antigen exposures but then subsequent evidence of waning (and diminished protection from infection six months after vaccination) informed the decision to introduce booster doses in the UK and globally. A greater immune response following longer intervals between vaccine doses has also been reported by others, including T-cell and antibody responses^6,21–23^. Together the evidence supports the decision made in the UK, and subsequently other countries, to delay the interval between doses^27,28^. Our data has also shown a similar effect, with higher anti-S titres with extended intervals between infection and vaccination. It is important to recognise however that within the SIREN cohort we have not observed a corresponding difference in protection against SARS-CoV-2 infection associated with timing of dosing interval^13^. Our data also adds to the literature on lower vaccine responses in older age groups^29–32^ and individuals with immunosuppression^33,34^, which should inform continued targeted interventions for these groups. Furthermore, with participants having diverse infection history (Wuhan, Alpha, Delta) we are now looking at the role of prior infection and antigenic imprinting on vaccination responses^35^.

Whilst this analysis involved a large population, enabling us to investigate several explanatory and control for important confounders, as a cross-sectional design, we have not followed-up individuals longitudinally. This means we are unable to assess the durability of antibody response in this analysis. Our data is also limited to anti-S titres and does not provide data on neutralising antibody response^36^ or T-cells responses^22^ as others have shown. It is also important to recognise that we were unable to investigate important factors such as ethnicity and vaccine type in this analysis given the SIREN cohort is predominately of white ethnicity and received the BNT162b2 vaccine. A longitudinal analysis including neutralising antibody testing is underway investigating responses to booster vaccine doses.

The SIREN cohort continues to provide insights into vaccine effectiveness, vaccine responses, reinfections and now vaccination breakthrough infections. With the waning of immunity and increasing vaccine breakthroughs, this cohort will play an essential part in determining why patients develop vaccine breakthrough infections and determining any potential correlates of protection.

## Conclusion

The SIREN cohort continues to provide insights into vaccine effectiveness, vaccine responses, reinfections and now vaccination breakthrough infections. With the waning of immunity and increasing vaccine breakthroughs, this cohort will play an essential part in determining why patients develop vaccine breakthrough infections and determining any potential correlates of protection.

## Data Availability

Annotated code for this analysis is available at: (https://github.com/SIREN-study/SARS-CoV-2-Immunity). The metadata for this analysis will be available to researchers through the Health Data Research UK CO-CONNECT platform and available for secondary analysis.

## Acknowledgments

We thank all SIREN participants for enrolling within the SIREN study and continuing to provide samples. We also thank all staff within the NHS that enable SIREN to operate.

Additionally, we thank the staff and students at UKHSA Porton Down that conducted the sample receipt and testing of baseline SIREN samples.

## Author contributions

ADO, VH, SH, HW, TB and AS conceptualised this analysis. ADO wrote the initial manuscript, with input from AS, VH, SF, SH, TB, HW, JH, SD’A and AA. ADO, SD’A, JH, CR, AS and TB oversaw the serological testing. EL and ST are responsible for sample collation, shipping and archiving, with support from JH. HW supported statistical analysis. VH and SH design and lead on the SIREN study. MC, EL, ST, AT-K, NH, NS, DC, IM, CT, CSB and JI support sample receipt, movement of samples and general SIREN operations.

## Ethics

This study was registered, number ISRCTN11041050, and received approval from the Berkshire Research Ethics Committee on 22 May 2020

