## Supplementary material for "Determinants of SARS-CoV-2 anti-spike antibody levels following BNT162b2 vaccination: cross-sectional analysis of 6,000 SIREN study participants"

### SIREN Study group

| No. | Institution | First name | Surname |
|---|---|---|---|
| 1. | UK Health Security Agency | Ana | Atti |
| 2. | UK Health Security Agency | Omoyeni | Adebiyi |
| 3. | UK Health Security Agency | Nick | Andrews |
| 4. | UK Health Security Agency | Tim | Brooks |
| 5. | UK Health Security Agency | Colin | Brown |
| 6. | UK Health Security Agency | Davina | Calbraith |
| 7. | UK Health Security Agency | Meera | Chand |
| 8. | UK Health Security Agency | Andre | Charlett |
| 9. | UK Health Security Agency | Michelle | Cole |
| 10. | UK Health Security Agency | Tom | Coleman |
| 11. | UK Health Security Agency | Joanna | Conneely |
| 12. | UK Health Security Agency | Paul | Conneely |
| 13. | UK Health Security Agency | Eleanor | Cross |
| 14. | UK Health Security Agency | Silvia | D’Arcangelo |
| 15. | UK Health Security Agency | Sarah | Foulkes |
| 16. | UK Health Security Agency | Nabila | Fowles-Gutierrez |
| 17. | UK Health Security Agency | Eileen | Gallagher |
| 18. | UK Health Security Agency | Natalie | Gillson |
| 19. | UK Health Security Agency | Victoria | Hall |
| 20. | UK Health Security Agency | Nipu | Hettiarachchi |
| 21. | UK Health Security Agency | Jacqueline | Hewson |
| 22. | UK Health Security Agency | Bethany | Hicks |
| 23. | UK Health Security Agency | Susan | Hopkins |
| 24. | UK Health Security Agency | Kate | Howell |
| 25. | UK Health Security Agency | Ferdinando | Insalata |
| 26. | UK Health Security Agency | Jasmin | Islam |
| 27. | UK Health Security Agency | Jameel | Khawam |
| 28. | UK Health Security Agency | Robert | Kyffin |
| 29. | UK Health Security Agency | Ezra | Linley |
| 30. | UK Health Security Agency | Iain | Milligan |
| 31. | UK Health Security Agency | Sebastian | Milward |
| 32. | UK Health Security Agency | Edward | Monk |
| 33. | UK Health Security Agency | Katie | Munro |
| 34. | UK Health Security Agency | Claire | Neill |
| 35. | UK Health Security Agency | Anne-Marie | O’Connell |
| 36. | UK Health Security Agency | Ashley | Otter |
| 37. | UK Health Security Agency | Mary | Ramsay |
| 38. | UK Health Security Agency | Cathy | Rowe |
| 39. | UK Health Security Agency | Ayoub | Saei |
| 40. | UK Health Security Agency | Noshin | Sajedi |
| 41. | UK Health Security Agency | Amanda | Semper |
| 42. | UK Health Security Agency | Andrew | Taylor-Kerr |
| 43. | UK Health Security Agency | Yrene | Themistocleous |
| 44. | UK Health Security Agency | Jean | Timeyin |
| 45. | UK Health Security Agency | Simon | Tonge |
| 46. | UK Health Security Agency | Caio | Tranquillini |
| 47. | UK Health Security Agency | Edgar | Wellington |
| 48. | UK Health Security Agency | Maria | Zambon |
| 49. | Public Health Agency Northern Ireland | Dianne | Corrigan |
| 50. | Public Health Agency Northern Ireland | Lisa | Cromey |

| | | | |
|---|---|---|---|
| 51. | Glasgow Caledonian University & Public Health Scotland | Lesley | Price |
| 52. | Glasgow Caledonian University & Public Health Scotland | Sally | Stewart |
| 53. | Glasgow Caledonian University & Public Health Scotland | Nicola | Sergenson |
| 54. | Public Health Scotland | Jennifer | Bishop |
| 55. | Public Health Scotland | Jennifer | Weir |
| 56. | Glasgow Caledonian University | Ayo | Matuluko |
| 57. | Glasgow Caledonian University | Annelysse | Jorgenson |
| 58. | Public Health Scotland | Laura | Dobbie |
| 59. | Public Health Scotland | Andrew | Telfer |
| 60. | Public Health Scotland | David | Goldberg |
| 61. | Public Health Wales | Ellen | De Lacy |
| 62. | Public Health Wales | Guy | Stevens |
| 63. | Public Health Wales | Susannah | Froude |
| 64. | Public Health Wales | Linda | Tyson |
| 65. | Health and Care Research Wales | Yvette | Ellis |
| 66. | Health and Care Research Wales | Chris | Norman |
| **No.** | **Participating SIREN Sites** | **First name** | **Surname** |
| 1 | ALDER HEY CHILDREN'S NHS FOUNDATION TRUST | B | Larru |
| | | S | Mcwilliam |
| 2 | Aneurin Bevan University LHB | John | Northfield |
| | | Sean | Cutler |
| 3 | ASHFORD AND ST PETER'S HOSPITALS NHS FOUNDATION TRUST | Stephen | Winchester |
| | | Samuel | Rowley |
| 4 | BASILDON AND THURROCK UNIVERSITY HOSPITALS NHS FOUNDATION TRUST | Stacey | Pepper |
| | | Georgina | Butt |
| 5 | BEDFORDSHIRE HOSPITALS NHS FOUNDATION TRUST | Simantee | Guha |
| | | Philippa | Bakker |
| 6 | Belfast Health & Social Care Trust | Clodagh | Loughrey |
| | | A | Watt |
| 7 | Betsi Cadwaladr University LHB | Christian | Subbe |
| | | Alice | Thomas |
| 8 | BIRMINGHAM AND SOLIHULL MENTAL HEALTH NHS FOUNDATION TRUST | Manny | Bagary |
| | | Di | Baines |
| 9 | BIRMINGHAM COMMUNITY HEALTHCARE NHS FOUNDATION TRUST | Rebecca | Chapman |
| | | Lucy Booth | Booth |
| 10 | BLACK COUNTRY HEALTHCARE NHS FOUNDATION TRUST | Alison | Grant |
| | | Rebecca | Temple-Purcell |
| 11 | BLACKPOOL TEACHING HOSPITALS NHS FOUNDATION TRUST | Joanne | Howard |
| | | Emma | Ward |
| 12 | BOLTON NHS FOUNDATION TRUST | Chinari | Subudhi |
| 13 | BRIGHTON AND SUSSEX UNIVERSITY HOSPITALS NHS TRUST | Marion | Campbell |
| | | Andrew | Bexley |
| 14 | BUCKINGHAMSHIRE HEALTHCARE NHS TRUST | R | Penn |
| | | N | Wong |
| 15 | | G | Boyd |

| | | | |
|---|---|---|---|
| | CALDERDALE AND HUDDERSFIELD NHS FOUNDATION TRUST | A | Rajgopal |
| 16 | CENTRAL AND NORTH WEST LONDON NHS FOUNDATION TRUST | Abigail | Severn |
| | | R | Matthews |
| 17 | CHESTERFIELD ROYAL HOSPITAL NHS FOUNDATION TRUST | Edward | Harris |
| | | Amanda | Whileman |
| 18 | CORNWALL PARTNERSHIP NHS FOUNDATION TRUST | Richard | Laugharne |
| | | Joanna | Ledger |
| 19 | COUNTESS OF CHESTER HOSPITAL NHS FOUNDATION TRUST | T | Barnes |
| | | C | Jones |
| 20 | CROYDON HEALTH SERVICES NHS TRUST | Banerjee | SubhroOsuji |
| | | Anna | Rokakis |
| 21 | Cwm Taf Morgannwg University LHB | John | Geen |
| | | Carla | Pothecary |
| 22 | DARTFORD AND GRAVESHAM NHS TRUST | Tracy | Edmunds |
| | | Nihil | Chitalia |
| 23 | DERBYSHIRE COMMUNITY HEALTH SERVICES NHS FOUNDATION TRUST | Sarah | Creer |
| | | Eve | Etell Kirby |
| 24 | DERBYSHIRE HEALTHCARE NHS FOUNDATION TRUST | S | Akhtar |
| | | G | Harrison |
| 25 | DEVON PARTNERSHIP NHS TRUST | Clare | McAdam |
| | | Natalie | Crooks |
| 26 | DONCASTER AND BASSETLAW TEACHING HOSPITALS NHS FOUNDATION TRUST | K | Agwuh |
| | | V | Maxwell |
| 27 | DORSET COUNTY HOSPITAL NHS FOUNDATION TRUST | Jennifer | Graves |
| 28 | DORSET HEALTHCARE UNIVERSITY NHS FOUNDATION TRUST | James | Colton |
| 29 | EAST SUFFOLK AND NORTH ESSEX NHS FOUNDATION TRUST | A | O'Kelly |
| | | P | Ridley |
| 30 | EAST SUSSEX HEALTHCARE NHS TRUST | Anna | Cowley |
| | | Janet | Sinclair |
| 31 | EPSOM AND ST HELIER UNIVERSITY HOSPITALS NHS TRUST | Helen | Johnstone |
| | | Neringa | Vilimiene |
| 32 | FRIMLEY HEALTH NHS FOUNDATION TRUST | Manjula | Meda |
| | | Jane | Democratis |
| 33 | GEORGE ELIOT HOSPITAL NHS TRUST | Simon | Brake |
| | | David | Boss |
| 34 | GLOUCESTERSHIRE HOSPITALS NHS FOUNDATION TRUST | Amanda | Selassie |
| | | Rekha Prince | Plackal |
| 35 | Golden Jubilee National Hospital | Catherine | Sinclair |
| | | Val | Irvine |
| 36 | GREAT WESTERN HOSPITALS NHS FOUNDATION TRUST | Eva | Fraile |
| 37 | HAMPSHIRE HOSPITALS NHS FOUNDATION TRUST | Claire | Thomas |
| | | Ina | Hoad |
| 38 | | Shekoo | Mackay |

| | | | |
|---|---|---|---|
| | HOUNSLOW AND RICHMOND COMMUNITY HEALTHCARE NHS TRUST | Shivani | Khan |
| 39 | HULL UNIVERSITY TEACHING HOSPITALS NHS TRUST | Philippa | Burns |
| | | Nicholas | Easom |
| 40 | Hywel Dda University LHB | Tracy | Lewis |
| | | Zohra | Omar |
| 41 | IMPERIAL COLLEGE HEALTHCARE NHS TRUST | Graham | Pickard |
| | | Kenisha | Lewis |
| 42 | ISLE OF WIGHT NHS TRUST | Sarah | Hinch |
| | | Alison | Brown |
| 43 | JAMES PAGET UNIVERSITY HOSPITALS NHS FOUNDATION TRUST | Ben | Burton |
| | | Christian | Hacon |
| 44 | KING'S COLLEGE HOSPITAL NHS FOUNDATION TRUST | Ray | Chaudhuri |
| | | Jonnie | Aeron-Thomas |
| 45 | LANCASHIRE & SOUTH CUMBRIA NHS FOUNDATION TRUST | Robert | Shorten |
| | | Kathryn | Williams |
| 46 | LANCASHIRE TEACHING HOSPITALS NHS FOUNDATION TRUST | Maya | Leach |
| | | Robert | Shorten |
| 47 | LEEDS TEACHING HOSPITALS NHS TRUST | Kyra | Holliday |
| | | Clair | Favager |
| 48 | LEICESTERSHIRE PARTNERSHIP NHS TRUST | Sarah | Baillon |
| | | Samantha | Hamer |
| 49 | LEWISHAM AND GREENWICH NHS TRUST | A | Shah |
| | | J | Russell |
| 50 | LINCOLNSHIRE PARTNERSHIP NHS FOUNDATION TRUST | Kelly | Moran |
| | | Ananta | Dave |
| 51 | LIVERPOOL UNIVERSITY HOSPITALS NHS FOUNDATION TRUST | Anu | Chawla |
| | | Fran | Westwell |
| 52 | LONDON NORTH WEST UNIVERSITY HEALTHCARE NHS TRUST | Ekaterina | Watson |
| | | D | Adeboyeku |
| 53 | MAIDSTONE AND TUNBRIDGE WELLS NHS TRUST | C | Pegg |
| | | M | Williams |
| 54 | MANCHESTER UNIVERSITY NHS FOUNDATION TRUST | S | Ahmad |
| | | A | Horsley |
| 55 | MID CHESHIRE HOSPITALS NHS FOUNDATION TRUST | Claire | Gabriel |
| | | Katherin | Pagett |
| 56 | MID ESSEX HOSPITAL SERVICES NHS TRUST | Lauren | Sach |
| | | Yvonne | Lester |
| 57 | MID YORKSHIRE HOSPITALS NHS TRUST | Ismaelette | Del Rosario |
| | | John | Ashcroft |
| 58 | MOORFIELDS EYE HOSPITAL NHS FOUNDATION TRUST | Roxanne | Crosby-Nwaobi |
| | | Chloe | Reeks |
| 59 | NHS Borders | Joy | Dawson |
| | | Lauren | Finlayson |
| 60 | NHS Fife | Susan | Fowler |
| | | Devesh | Dhasmana |

| | | | |
|---|---|---|---|
| 61 | NHS Forth Valley | Euan | Cameron |
| | | Anne | Todd |
| 62 | NHS Grampian | Harriet | Carroll |
| | | Alison | Thornton |
| 63 | NHS Greater Glasgow and Clyde | Antonia | Ho |
| | | Michael | Murphy |
| 64 | NHS Highland | Andrew | Gibson |
| | | Alexandra | Cochrane |
| 65 | NHS Lanarkshire | Manish | Patel |
| | | Karen | Black |
| 66 | NHS Lothian | Kate | Templeton |
| | | Andrea | Clarke |
| 67 | NHS Western Isles | Martin | Malcolm |
| | | Joan | Frieslick |
| 68 | NORFOLK AND NORWICH UNIVERSITY HOSPITALS NHS FOUNDATION TRUST | Ngozi | Elumogo |
| | | Louise | Coke |
| 69 | NORTH CUMBRIA INTEGRATED CARE NHS FOUNDATION TRUST | Beverly | Wilkinson |
| | | John | Elliott |
| 70 | NORTH MIDDLESEX UNIVERSITY HOSPITAL NHS TRUST | Mariyam | Mirfenderesky |
| | | Pratap | Harbham |
| 71 | NORTH WEST ANGLIA NHS FOUNDATION TRUST | Janki | Bhayani |
| | | Stephanie | Diaz |
| 72 | NORTHERN DEVON HEALTHCARE NHS TRUST | M | Howard |
| | | T | Lewis |
| 73 | NORTHERN HEALTH AND SOCIAL CARE TRUST | Elinor | Hanna |
| | | Frances | Johnston |
| 74 | NORTHERN LINCOLNSHIRE AND GOOLE NHS FOUNDATION TRUST | Jonathan | Hatton |
| | | Peter | Cowling |
| 75 | NOTTINGHAM UNIVERSITY HOSPITALS NHS TRUST | Sarah | Brand |
| | | Imogen | Gould |
| 76 | POOLE HOSPITAL NHS FOUNDATION TRUST | Beverley | Wadams |
| | | Elizabeth | Sheridan |
| 77 | PORTSMOUTH HOSPITALS NHS TRUST | Johanna | Mouland |
| | | Jade | Yates |
| 78 | Powys Teaching LHB | Jayne | Goodwin |
| | | Chris | Norman |
| 79 | QUEEN VICTORIA HOSPITAL NHS FOUNDATION TRUST | J | Giles |
| | | G | Pottinger |
| 80 | ROYAL BERKSHIRE NHS FOUNDATION TRUST | Maya | Joseph |
| | | Holly | Coles |
| 81 | ROYAL CORNWALL HOSPITALS NHS TRUST | H | Chenoweth |
| | | D | Browne |
| 82 | ROYAL DEVON AND EXETER NHS FOUNDATION TRUST | Cressida | Auckland |
| | | Stephanie | Prince |
| 83 | | Alison | Rodger |

| | | | |
|---|---|---|---|
| | ROYAL FREE LONDON NHS FOUNDATION TRUST | Tabitha | Mahungu |
| 84 | ROYAL NATIONAL ORTHOPAEDIC HOSPITAL NHS TRUST | Esther | Hanison |
| | | Simon | Warren |
| 85 | ROYAL PAPWORTH HOSPITAL NHS FOUNDATION TRUST | Sumita | Pai |
| | | Helen | Baxendale |
| 86 | ROYAL SURREY COUNTY HOSPITAL NHS FOUNDATION TRUST | Charles | Piercy |
| | | Esther | Tarr |
| 87 | ROYAL UNITED HOSPITALS BATH NHS FOUNDATION TRUST | Debbie | Delgado |
| | | Sarah | Meisner |
| 88 | SALISBURY NHS FOUNDATION TRUST | Catherine | Thompson |
| | | Sophia | Strong-Sheldrake |
| 89 | SANDWELL AND WEST BIRMINGHAM HOSPITALS NHS TRUST | Ash | Turner |
| | | Anne | Hayes |
| 90 | SHEFFIELD CHILDREN'S NHS FOUNDATION TRUST | S | Gormley |
| | | C | Kerrison |
| 91 | SHEFFIELD TEACHING HOSPITALS NHS FOUNDATION TRUST | Thushan | de Silva |
| | | Simon | Tazzyman |
| 92 | SHERWOOD FOREST HOSPITALS NHS FOUNDATION TRUST | Lynne | Allsop |
| | | Shrikant | Ambalkar |
| 93 | SHREWSBURY AND TELFORD HOSPITAL NHS TRUST | Mandy | Beekes |
| | | Hannah | Gibson |
| 94 | SHROPSHIRE COMMUNITY HEALTH NHS TRUST | Johanne | Tomlinson |
| 95 | SOLENT NHS TRUST | Cathy | Price |
| | | The Solent Research Team | |
| 96 | SOMERSET NHS FOUNDATION TRUST | Justin | Pepperell |
| | | Kate | James |
| 97 | South Eastern Health & Social Care | Yuri | Protaschik |
| | South Eastern Health & Social Care | Tom | Trinick |
| 98 | SOUTHEND UNIVERSITY HOSPITAL NHS FOUNDATION TRUST | John | Day |
| | | Swapna | Kunhunny |
| 99 | Southern Health & Social Care Trust | Angel | Boulos |
| | | Alice | Neave |
| 100 | SOUTHERN HEALTH NHS FOUNDATION TRUST | Qi | Zheng |
| 101 | SOUTHPORT AND ORMSKIRK HOSPITAL NHS TRUST | Katherine | Gray |
| | | Kerryanne | Brown |
| 102 | ST GEORGE'S UNIVERSITY HOSPITALS NHS FOUNDATION TRUST | Tim | Planche |
| | | Angela | Houston |
| 103 | ST HELENS AND KNOWSLEY TEACHING HOSPITALS NHS TRUST | Rowan | Pritchard Jones |
| | | Diane | Wycherley |
| 104 | STOCKPORT NHS FOUNDATION TRUST | Barzo | Faris |
| 105 | SURREY AND SUSSEX HEALTHCARE NHS TRUST | K | Nimako |
| | | B | Stewart |

| | | | |
|---|---|---|---|
| 106 | Swansea Bay University LHB | Claire | Stafford |
| | | Rebeccah | Thomas |
| 107 | THE CLATTERBRIDGE CANCER CENTRE NHS FOUNDATION TRUST | Sheena | Khanduri |
| | | Nagesh | Kalakonda |
| 108 | THE DUDLEY GROUP NHS FOUNDATION TRUST | Helen | Ashby |
| 109 | THE HILLINGDON HOSPITALS NHS FOUNDATION TRUST | Natasha | Mahabir |
| 110 | THE NEWCASTLE UPON TYNE HOSPITALS NHS FOUNDATION TRUST | J | Harwood |
| | | B | Payne |
| 111 | THE PRINCESS ALEXANDRA HOSPITAL NHS TRUST | Kathryn | Court |
| | | Nikki | White |
| 112 | THE ROBERT JONES AND AGNES HUNT ORTHOPAEDIC HOSPITAL NHS FOUNDATION TRUST | Ruth | Longfellow |
| 113 | THE ROYAL BOURNEMOUTH AND CHRISTCHURCH HOSPITALS NHS FOUNDATION TRUST | Mihye | Lee |
| 114 | THE ROYAL WOLVERHAMPTON NHS TRUST | Marie | Green |
| | | Lauren | Hughes |
| 115 | TORBAY AND SOUTH DEVON NHS FOUNDATION TRUST | Mathew | Halkes |
| | | Pauline | Mercer |
| 116 | UNITED LINCOLNSHIRE HOSPITALS NHS TRUST | Alun | Roebuck |
| | | | ULHT Research Team |
| 117 | UNIVERSITY HOSPITAL SOUTHAMPTON NHS FOUNDATION TRUST | E | Wilson-Davies |
| 118 | UNIVERSITY HOSPITALS BRISTOL AND WESTON NHS FOUNDATION TRUST | Rajeka | Lazarus |
| | | Aaran | Sinclair |
| 119 | UNIVERSITY HOSPITALS COVENTRY AND WARWICKSHIRE NHS TRUST | N | Aldridge |
| | | L | Berry |
| 120 | UNIVERSITY HOSPITALS OF DERBY AND BURTON NHS FOUNDATION TRUST | F | Game |
| | | T | Reynolds |
| 121 | UNIVERSITY HOSPITALS OF LEICESTER NHS TRUST | Christopher | Holmes |
| | | Martin | Wiselka |
| 122 | UNIVERSITY HOSPITALS OF MORECAMBE BAY NHS FOUNDATION TRUST | Lynda | Fothergill |
| | | Karen | Burns |
| 123 | UNIVERSITY HOSPITALS OF NORTH MIDLANDS NHS TRUST | Christopher | Duff |
| | | Martin | Booth |
| 124 | UNIVERSITY HOSPITALS PLYMOUTH NHS TRUST | Hannah | Jory |
| | | David | Hilton |
| 125 | Velindre NHS Trust | Charlotte | Young |
| | | James | Powell |
| 126 | WALSALL HEALTHCARE NHS TRUST | Lisa | Richardson |
| | | Aiden | Plant |
| 127 | | Zaman | Qazzafi |

| | | | |
|---|---|---|---|
| | WARRINGTON AND HALTON TEACHING HOSPITALS NHS FOUNDATION TRUST | Lisa | Ditchfield |
| 128 | WEST SUFFOLK NHS FOUNDATION TRUST | A | Moody |
| | | R | Tilley |
| 129 | Western Health & Social Care Trust | Tracy | Donaghy |
| | | Maurice | O'Kane |
| 130 | WESTERN SUSSEX HOSPITALS NHS FOUNDATION TRUST | R | Sierra |
| | | K | Shipman |
| 131 | WHITTINGTON HEALTH NHS TRUST | Philippa | Kemsley |
| | | Chetan | Parmar |
| 132 | WIRRAL UNIVERSITY TEACHING HOSPITAL NHS FOUNDATION TRUST | D | Harvey |
| | | Y | Huang |
| 133 | WYE VALLEY NHS TRUST | L | Robinson |
| 134 | YEOVIL DISTRICT HOSPITAL NHS FOUNDATION TRUST | Sarah | Board |
| | | Andrew | Broadley |
| 135 | YORK TEACHING HOSPITAL NHS FOUNDATION TRUST | Claire | Brookes |
| | | Mags | Szewczyk |
| **No.** | **SIREN Associated Studies** | **First name** | **Surname** |
| 1. | Protective Immunity from T cells to Covid-19 in Health workers (PITCH) | Susanna | Dunachie |
| 2. | Protective Immunity from T cells to Covid-19 in Health workers (PITCH) | Paul | Klenerman |
| 3. | Protective Immunity from T cells to Covid-19 in Health workers (PITCH) | Chris | Duncan |
| 4. | Protective Immunity from T cells to Covid-19 in Health workers (PITCH) | Lance | Turtle |
| 5. | Protective Immunity from T cells to Covid-19 in Health workers (PITCH) | Alex | Richter |
| 6. | Protective Immunity from T cells to Covid-19 in Health workers (PITCH) | Thushan | De Silva |
| 7. | Protective Immunity from T cells to Covid-19 in Health workers (PITCH) | Eleanor | Barnes |
| 8. | Protective Immunity from T cells to Covid-19 in Health workers (PITCH) | Daniel | Wootton |
| 9. | The Humoral Immune Correlates for COVID-19 (HICC) consortium | Jonathan | Heeney |
| 10. | The Humoral Immune Correlates for COVID-19 (HICC) consortium | Helen | Baxendale |
| 11. | The Humoral Immune Correlates for COVID-19 (HICC) consortium | Javier | Castillo-Olivares |
| 12. | The Francis Crick Institute | Rupert | Beale |
| 13. | The Francis Crick Institute | Edward | Carr |
| 14. | Genotype2Phenotype (G2P) | Wendy | Barclay |
| 15. | Genotype2Phenotype (G2P) | Massimo | Palmarini |
| 16. | GenOMICC | John Kenneth | Baillie |

### Supplementary tables

**Supplementary table 1:** Overview of the post-infection and post-vaccination cohort, split according to gender and ethnic group. Median (and min/max) age is also given for each gender group.

| Group | Gender | n= | Median age (range) | Ethnic group | n= |
|---|---|---|---|---|---|
| Post-infection, pre-vaccination | Male | 1297 | 4 (19 – 76) | White | 937 |
| | | | | Black | 358 |
| | | | | Other/prefer not to say | 2 |
| | Female | 5824 | 46 (19 – 78) | White | 5013 |
| | | | | Black | 802 |
| | | | | Other/prefer not to say | 9 |
| | Other/prefer not to say | 10 | - | - | - |
| Post dose 1 | Male | 640 | 43 (18 – 70) | White | 517 |
| | | | | Black | 122 |
| | | | | Other/prefer not to say | 1 |
| | Female | 3344 | 46 (19 – 76) | White | 3134 |
| | | | | Black | 206 |
| | | | | Other/prefer not to say | 4 |
| | Other/prefer not to say | 5 | - | - | - |
| Post dose 2 | Male | 354 | 45 (19 – 73) | White | 263 |
| | | | | Black | 90 |
| | | | | Other/prefer not to say | 1 |
| | Female | 1526 | 47 (20 – 69) | White | 1427 |
| | | | | Black | 99 |
| | Other/prefer not to say | 2 | - | - | - |

**Supplementary table 2:** Multivariable regression model used within this study, with groups and subgroups for analysis.

| Group | Subgroup | Adjusted geometric mean ratio (95% CI) | P>Z |
|---|---|---|---|
| Dose 1, uninfected, >28 days | | | |
| Age | 18-24 | 2.28 (1.74 - 2.99) | <0.0001 |
| | 25-34 | 1.99 (1.74 - 2.27) | <0.0001 |
| | 35-44 | 1.46 (1.29 - 1.65) | <0.0001 |
| | 45-54 | 1.19 (1.05 - 1.34) | 0.005 |
| | 55+ | 1 (ref) | - |
| Gender | Male | 1 (ref) | - |
| | Female | 1.25 (1.11 - 1.41) | <0.0001 |
| | Other/Unknown | 1.11 (0.31 - 3.97) | 0.872 |
| Ethnicity | White | 1 (ref) | - |
| | BAME | 1.14 (0.96 - 1.35) | 0.147 |
| | Other/Unknown | 0.77 (0.16 - 3.61) | 0.735 |
| Condition | none | 1 (ref) | - |
| | Immunocompromised | 0.61 (0.46 - 0.81) | 0.001 |
| | Chronic Respiratory conditions | 0.96 (0.84 - 1.1) | 0.566 |
| | Chronic non-Respiratory conditions | 0.81 (0.71 - 0.94) | 0.005 |
| Dose 2, uninfected, >21 days | | | |
| Age | 18-24 | 2.34 (1.45 - 3.78) | 0.001 |
| | 25-34 | 1.65 (1.36 - 2) | <0.0001 |
| | 35-44 | 1.44 (1.22 - 1.7) | <0.0001 |
| | 45-54 | 1.23 (1.04 - 1.44) | 0.013 |
| | 55+ | 1 (ref) | 0.001 |
| Gender | Male | 1 (ref) | |
| | Female | 1.12 (0.96 - 1.32) | 0.151 |
| | Other/Unknown | 2.27 (0.33 - 15.39) | 0.402 |
| Ethnicity | White | 1 (ref) | |
| | BAME | 1.27 (1.03 - 1.55) | 0.024 |
| | Other/Unknown | 0.91 (0.13 - 6.12) | 0.92 |
| Condition | none | 1 (ref) | - |
| | Immunocompromised | 0.38 (0.27 - 0.55) | <0.0001 |
| | Chronic Respiratory conditions | 1.06 (0.89 - 1.27) | 0.512 |
| | Chronic non-Respiratory conditions | 1 (0.83 - 1.21) | 0.994 |
| Dose interval | 2-3 weeks | 0.19 (0.15 - 0.25) | <0.0001 |
| | 4-6 weeks | 0.35 (0.3 - 0.42) | <0.0001 |
| | 7-9 weeks | 0.73 (0.62 - 0.86) | <0.0001 |
| | 10-12+ weeks | 1 (ref) | - |

| Dose 2, previously infected, >21 days | | | |
|---|---|---|---|
| Age | 18-24 | 1.42 (0.75 - 2.7) | 0.28 |
| | 25-34 | 0.84 (0.57 - 1.24) | 0.381 |
| | 35-44 | 0.88 (0.59 - 1.29) | 0.504 |
| | 45-54 | 0.9 (0.63 - 1.27) | 0.54 |
| | 55+ | 1 (ref) | - |
| Gender | Male | 1 (ref) | - |
| | Female | 0.77 (0.56 - 1.05) | 0.099 |
| Ethnicity | White | 1 (ref) | - |
| | BAME | 1.17 (0.8 - 1.69) | 0.421 |
| Condition | None | 1 (ref) | - |
| | Immunocompromised | 1.21 (0.63 - 2.34) | 0.563 |
| | Chronic Respiratory conditions | 0.96 (0.65 - 1.41) | 0.83 |
| | Chronic non-Respiratory conditions | 1.55 (1.04 - 2.32) | 0.032 |
| Dose interval | 2-3 weeks | 0.98 (0.54 - 1.78) | 0.94 |
| | 4-6 weeks | 1.07 (0.77 - 1.48) | 0.696 |
| | 7-9 weeks | 1.1 (0.79 - 1.54) | 0.574 |
| | 10-12+ weeks | 1 (ref) | - |

**Supplementary Table 3: Anti-spike antibody levels (BAU/ml) of naïve participants post-second vaccine dose, split according to dosing interval and time between 2nd dose and sample date.** Data are presented as geometric means, with 95% CIs. All groups show significantly higher Roche S values with greater dosing interval. Cohorts are split according to time between second vaccination and sample date.

| | | Time between 2nd dose and sample date | | | | | | | | | |
|---|---|---|---|---|---|---|---|---|---|---|---|
| | | 2-3 weeks | | 4-5 weeks | | 6-7 weeks | | 8-9 weeks | | 10+ weeks | |
| Dosing Interval | | n= | GMC (95% CI) | n= | GMC (95% CI) | n= | GMC (95% CI) | n= | GMC (95% CI) | n= | GMC (95% CI) |
| | **>2 and <4 weeks** | 4 | 1,657.14 (545.59 - 5033.25) | 18 | 1,855.3 (1237.02 - 2782.6) | 22 | 1,056.48 (703.21 - 1587.22) | 22 | 1,034.61 (729.33 - 1467.67) | 27 | 1,297.52 (820.08 - 2052.92) |
| | **>4 and <7 weeks** | 61 | 4,552.65 (3248.83 - 6379.71) | 94 | 3,788.21 (3143.54 - 4565.1) | 73 | 2,820.42 (2355.52 - 3377.07) | 36 | 1,828.95 (1199.77 - 2788.07) | 48 | 1,816.83 (1419.12 - 2326) |
| | **>7 and <10** | 114 | 11,811.87 (9766.09 - 14286.2) | 128 | 7,294 (5883.1 - 9043.28) | 63 | 5,325.2 (4405.78 - 6436.48) | 21 | 5,650.38 (4247.83 - 7516.01) | 2 | 4,304.73 (58.21 - 318355.14) |
| | **>10 weeks** | 398 | 12,985.38 (11906.36 - 14162.2) | 338 | 10,493.94 (9433.82 - 11673.18) | 50 | 7,927.22 (5916.64 - 10621.04) | 4 | 10,950.53 (2227.8 - 53826.19) | 0 | N/A |

### Supplementary figures

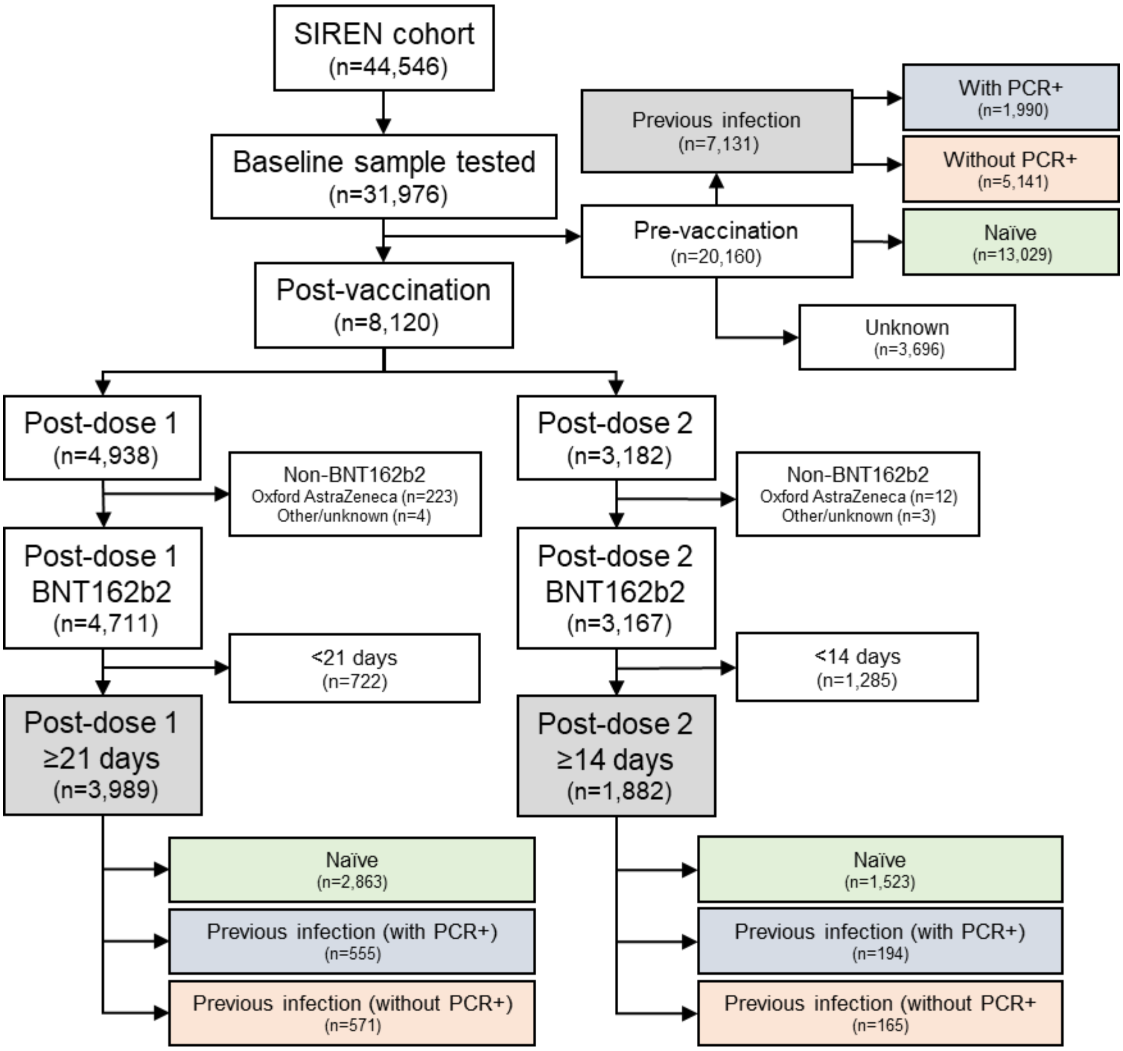


**Supplementary figure 1**: Inclusion and exclusion criteria used for the SIREN post-vaccination cross-sectional analysis. Boxes highlighted in grey were the final data sets used within this analysis, with further stratification into naïve individuals and those with previous infection with PCR+ and previous infection without PCR+.

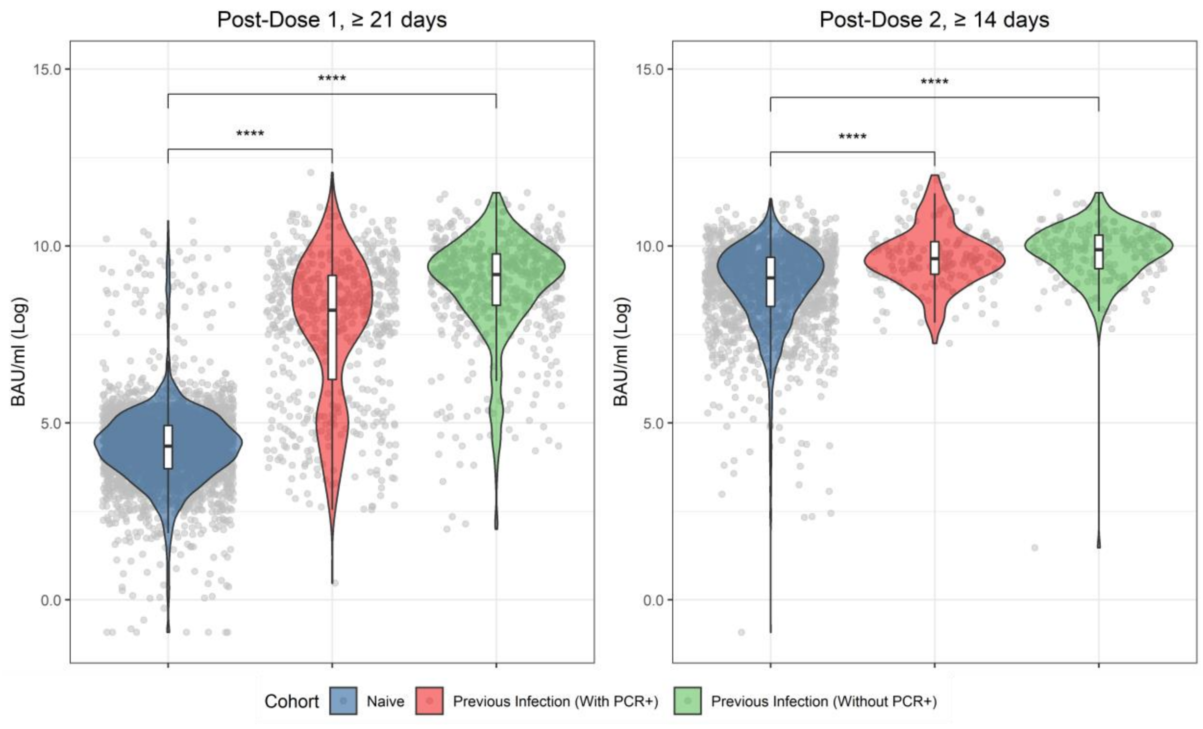


**Supplementary figure 2: Violin plot of anti-spike binding antibody levels (BAU/ml) post-dose 1 (left) and post-dose 2 (right), split by prior infection status.** Naive - those with no known previous infection (no PCR confirmation or anti-nucleocapsid antibodies at baseline). Previous infection with PCR+ – those with a known date at which they became infected. Previous infection without PCR+ – those with no known history of being PCR+ but presence of anti-nucleocapsid antibodies. Unpaired T-test was used to determine significance between groups.

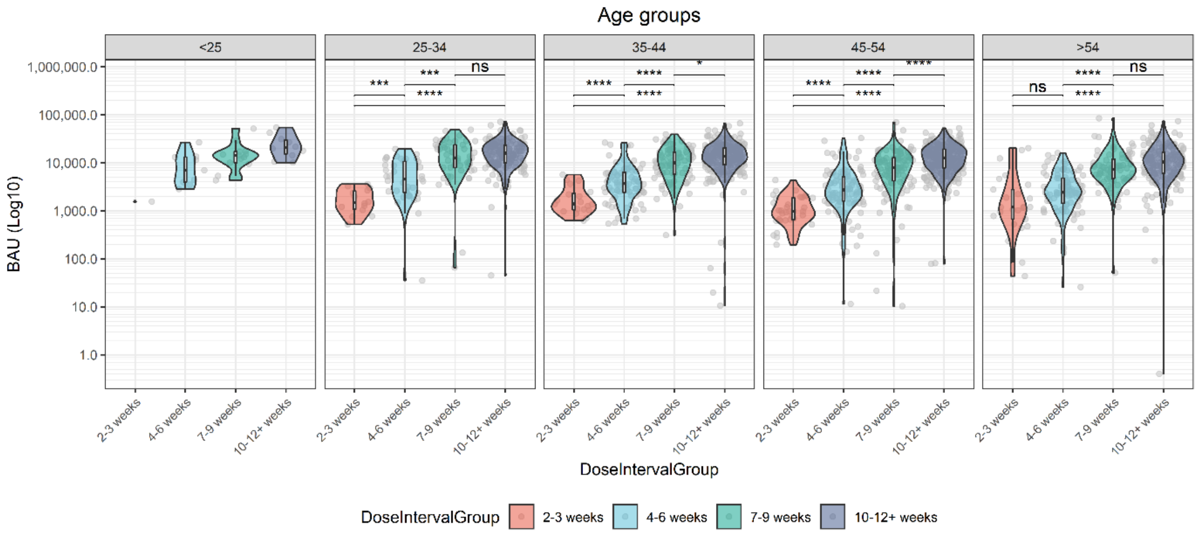


**Supplementary figure 3: Antibody levels post-dose 2 analysed according to dosing interval and age.** All age groups show a significantly higher antibody response (p=<0.0001) post-dose 2 where a 10-12-week dosing interval was used compared to a 2-3 week dosing interval. Unpaired T-test was used to determine significance between groups.

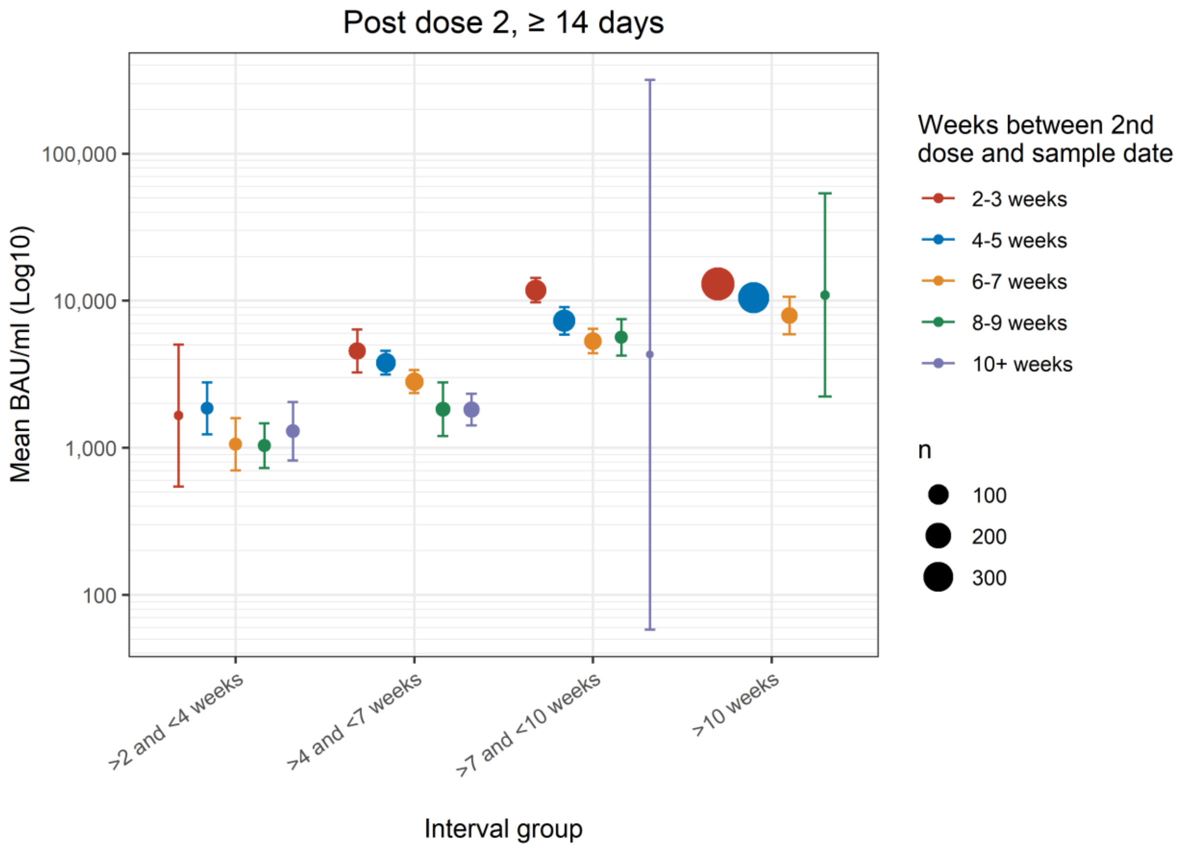


**Supplementary figure 4: Geometric mean anti-spike antibody levels (BAU/ml) of participants post-dose 2, split according to dosing interval and the time between second vaccination and sample collection**. Geometric mean antibody titres increase with increasing dosing interval, but antibody titres wane with an increase in weeks between sample date and date of second vaccination. Error bars shown are 95% CI.
